# Evaluating Accuracy and Reasoning Capabilities of Large Language Models for Acute Ischemic Stroke Management

**DOI:** 10.1101/2025.09.03.25335060

**Authors:** Aymen Meddeb, Navid Bakhtiari, Ida Rangus, Leonard Fetscher, Bastien Leguellec, Felix Busch, Alexandre Doucet, Vi-Tuan Hua, Fuong Verot-Nguyen, Laurentiu Valentin Paiusan, Pierre Francois Manceau, Paolo Pagano, Mike P. Wattjes, Solene Moulin, Laurent Pierot, Sebastien Soize

**Author notes:** **Corresponding Author:** Dr. Aymen Meddeb, Department of Neuroradiology, Hôpital Maison-Blanche, CHU Reims, Université Reims-Champagne-Ardenne, Reims, France.

## Abstract

**Background:** Acute ischemic stroke (AIS) management has evolved substantially over the past two decades, with mechanical thrombectomy adding complexity that requires specialized centers. As many patients initially present to primary care facilities, rapihtmld and accurate triage is critical. Large language models (LLMs) may help bridge expertise gaps, especially where stroke specialists are not immediately available. This study evaluates the diagnostic accuracy and reasoning quality of four LLMs in determining eligibility for intravenous thrombolysis (IVT) and mechanical thrombectomy (MT), compared with experienced clinicians and real-world treatment decisions.

**Methods:** We retrospectively collected 80 acute ischemic stroke cases from two stroke centers. Cases were presented to LLMs as well to clinicians as clinical vignettes containing demographic, clinical, and imaging data. Four LLMs (DeepSeek R1, OpenAI o3 mini, Gemini 2.0, LLaMA 3.3) and six stroke experts (two neurologists, four neuroradiologists) independently reviewed the cases and recommended one or more treatment strategies including IVT and MT. The ground truth was defined as the institutional treatment decision. Accuracy for MT and IVT recommendations was calculated for both LLMs and clinicians. Additionally, a qualitative error analysis evaluated the reasoning ability of LLMs.

**Results:** Open-source reasoning model DeepSeek R1 outperformed all other LLMs and clinicians for MT (87% accuracy) and achieved 78% accuracy for IVT. Across models, accuracy was generally higher for MT than for IVT. Neurologists reached 81% (MT) and 80% (IVT), while neuroradiologists achieved 84% (MT) and 76% (IVT). Reasoning analysis for MT recommendations showed that most errors were clinically reasonable but differed from real-world decision, whereas IVT errors were primarily due to guideline non-adherence.

**Conclusions:** LLMs can match or even exceed expert clinician performance in MT and IVT eligibility decisions, while providing transparent reasoning. These findings support prospective evaluation of LLM-based decision support in acute stroke care, especially in settings without immediate specialist expertise.

## Introduction

Since the landmark trials of 2015, mechanical thrombectomy (MT) has become the standard of care for selected patients with acute ischemic stroke due to large vessel occlusion (LVO) in the anterior circulation [1,2]. This therapeutic breakthrough has led to significant reductions in morbidity and mortality and has catalyzed national and international efforts to improve care for stroke patients [3]. Comprehensive stroke centers and tertiary hospitals have been at the forefront of these efforts, developing multidisciplinary stroke teams and training programs to ensure that stroke neurologists and interventional neuroradiologists are equipped with the expertise needed to deliver timely and effective reperfusion therapies [4]. Simultaneously, infrastructural investments have sought to optimize imaging workflows, streamline triage algorithms, and facilitate rapid patient transfers where needed [5].

However, a critical gap remains in the delivery of stroke care: patients presenting to non-tertiary or primary care centers are frequently assessed by physicians lacking expertise in stroke neurology or interventional neuroradiology [6]. In such settings, the decision whether or not to refer a patient for thrombectomy should be based on an interplay of clinical, radiological, and logistical factors, which may overwhelm non-specialists, particularly under time pressure [7,8]. This expertise gap leads to delays in reperfusion, inappropriate triage, or missed treatment opportunities, ultimately impacting patient outcomes [9]. Bridging this gap with scalable, accessible, and expert-level decision support is a pressing challenge in stroke systems of care.

Artificial intelligence, particularly through the emergence of large language models (LLMs), offers a promising pathway to addressing this challenge[10]. Beyond their initial applications in general language tasks, LLMs have rapidly found utility in medicine [11]. In neurology, LLMs have demonstrated impressive performance: passing board certification-style examinations [12], solving complex diagnostic cases, interpreting patient records [13], and even simulating empathetic physician-patient interactions to facilitate informed consent discussions [14]. In radiology, LLMs have been applied to generate structured report summaries [15], translate reports into multiple languages [16], extract key findings from unstructured clinical texts [17], and support diagnostic workflows [18].

Despite their growing relevance, the potential of LLMs as real-time clinical decision support tools in time-critical real-world scenarios such as acute stroke remains underexplored. Unlike diagnostic classification tasks or post hoc summarization of patient data, stroke management requires nuanced clinical reasoning, integration of multimodal data (e.g., clinical findings, imaging results, timing of symptom onset), and adherence to dynamic treatment guidelines [19].

In this study, we aim to evaluate the capabilities of state-of-the-art large language models in the triage and management of acute ischemic stroke patients. Specifically, we assess their performance in determining the eligibility for intravenous thrombolysis and mechanical thrombectomy across a cohort of real-world cases. We benchmark their decisions against two standards: (1) the real-world treatment decisions made by stroke teams in tertiary centers, and (2) decisions by experienced stroke neurologists and interventional neuroradiologists. Furthermore, we examine not only the accuracy of LLM recommendations but also the reasoning pathways and potential for clinical harm from incorrect suggestions. By systematically analyzing the strengths and limitations of LLMs in this domain, we aim to evaluate their readiness for deployment in clinical decision-making, especially in settings where expert knowledge is limited or absent.

## Methods

### Patient population

This study includes data from two independent datasets collected at two university hospitals located in two European countries, Charité – Medical University Berlin, Germany and Centre Hospitalier Universitaire de Reims, France, both certified as comprehensive stroke centers. The first dataset comprises reports from patients who underwent mechanical thrombectomy for acute ischemic stroke between September 2022 and June 2023 at the first institution. The second dataset includes consecutive patient reports from the second university hospital, covering the period from September 2023 to March 2024. All data collection procedures complied with the ethical principles of the Declaration of Helsinki. Beyond the patient’s age and sex, no identifiable features about the patient were transmitted to the LLM models; especially, no patient-identifying information was provided to the AI. This retrospective study received approval from the French National Research Ethics Committee for Medical Imaging (Reference: CRM-2501-444). The requirement for informed consent was waived due to the retrospective nature of the study.

### LLM Inference and Prompt Structure

Two commercial LLMs (OpenAI o3 mini and Gemini 2.0) and two open-source LLMs (LLaMA 3.3 and DeepSeek R1) were used in this study. The prompt was implemented using structured case vignettes derived from anonymized clinical reports. Each clinical vignette was directly incorporated into this template, followed by a clear and unambiguous question: *“Patient: [Age, Sex, Medical history, Medications, NIHSS score, Time since symptom onset, Imaging modality, ASPECTS score, Vessel occlusion, Collateral status]. Based on current acute ischemic stroke guidelines, should this patient receive Intravenous Thrombolysis (IVT), Mechanical Thrombectomy (MT), or a combination of both?”*

LLMs were instructed to provide both a recommendation and a brief justification. Inference was performed using the latest available checkpoints of o3 mini (OpenAI), LLaMA 3.3 (Meta), and Gemini 2.0 (Google), accessed through official APIs, and DeepSeek-R1 through Together AI platform.

To reduce randomness and ensure deterministic responses were possible, all LLM sessions were conducted with default model settings, with a temperature parameter set to 0. The very first response generated by each LLM for each vignette was recorded as its final therapeutic decision. No follow-up questions, clarifications, or feedback were provided to the models between cases.

### Human Rating

Six human experts participated in the rating task: two neurologists (I.R., A.D.) and four neuroradiologists (L.V.P., F.V.N, P.F.M, P.P), all with at least five years of experience in stroke care. Each expert independently reviewed the 80 anonymized vignettes, presented in identical order, and selected one or more appropriate treatments (IVT and/or MT). Responses were collected via Google Forms and subsequently encoded into binary variables for each treatment decision.

### Diagnostic Accuracy Evaluation

Final institutional decisions recorded in the clinical charts were defined as the ground truth (GT). For each rater (human or LLM), we computed diagnostic accuracy by comparing the binary predictions for each treatment category (IVT and MT) to the ground truth. Accuracy was calculated per individual and then averaged per group. Inter-rater agreement among human raters was assessed using Fleiss’ Kappa for each treatment and group. To complement accuracy metrics, we calculated the Jaccard index as a measure of agreement between predicted and ground truth treatment decisions. Unlike accuracy, which only captures correct versus incorrect classifications, the Jaccard index accounts for partial or complete overlap for multiple treatment categories.

### LLM Reasoning Evaluation

To assess model reasoning capabilities and identify failure modes, we conducted a structured qualitative error analysis of all false positive and false negative outputs from the four LLMs. Each incorrect LLM decision was independently reviewed by two authors (N.B. and A.M.) and categorized into one of four predefined error types:

- **Omission of critical information:** cases where the LLM failed to incorporate key details explicitly mentioned in the input.
- **Guideline non-adherence:** cases where the LLM’s decision contradicted published stroke treatment guidelines.
- **Borderline case misclassification:** cases involving nuanced scenarios with clinical ambiguity or gray zones, such as IVT eligibility beyond 4.5 hours or borderline ASPECTS scores.
- **Correct recommendation, discrepant ground truth:** cases where the LLM’s answer aligned with guidelines and input data but differed from the real-world decision, usually due to information that was unavailable to the LLM but known to clinicians.

Disagreements in categorization were resolved by consensus. We then quantified the distribution of these errors across models and treatment categories. Representative examples from each category were extracted to illustrate common reasoning patterns and limitations in model-generated recommendations.

## Results

### Patient population

The characteristics of the study cohort are shown in Table 1. The average age of study participants was 68.7 ± 14.7 years, with a near 1:1 sex ratio (51% of female patients, n=41). The patient population exhibited a high prevalence of established vascular risk factors consistent with typical stroke cohorts including hypertension (70%), diabetes mellitus (11%), and atrial fibrillation (10%). Approximately a quarter of the patients were on antiplatelet therapy (22%), and a lower proportion (17%) were treated with anticoagulants. The median NIHSS score on admission was 11 (Interquartile Range [IQR]: 5–16), indicating a patient population presenting with moderate to severe neurological deficits. MRI was performed as the initial neuroimaging modality in 50% of the cases (n = 40), using a standardized protocol that included diffusion-weighted imaging (DWI), fluid-attenuated inversion recovery (FLAIR), time-of-flight (TOF) angiography, and susceptibility-weighted imaging (SWI). 41% of the cases (n=33) received non-contrast CT plus CTA. Both imaging modalities were performed in six patients (7%). The mean ASPECTS score was 8.2 ± 2.1, suggesting limited early ischemic changes on baseline imaging for most patients.

**Table 1.** Characteristics of the Study Cohort (n=80)

| Characteristic | Value |
| --- | --- |
| Age, mean ± SD (years) | $68.7 \pm 14.7$ |
| Male sex, n (%) | 39 (49%) |
| Hypertension, n (%) | 56 (70%) |
| **Diabetes mellitus, n (%)** | 9 (11%) |
| **Atrial fibrillation, n (%)** | 8 (10%) |
| **Antiplatelet therapy, n (%)** | 18 (22%) |
| **Anticoagulant therapy, n (%)** | 14 (17%) |
| **NIHSS, median (IQR)** | 11 (5–16) |
| **Imaging: CT/MRI/both, n (%)** | 33 (41%) / 40 (50%) / 6 (7%) |
| **ASPECTS score, mean ± SD** | 8.2 ± 2.1 |

### Human Rating evaluation

Neurologists (n=2) achieved an accuracy of 81% for MT, and 80% for IVT. Neuroradiologists (n=4) achieved an accuracy of 84% for MT, and 76% for IVT. For MT eligibility, the interrater agreement was moderate across neurologists (κ = 0.618) and neuroradiologists (κ = 0.595). For IVT decisions, there was a substantial agreement among neurologists (κ = 0.725) but only fair agreement among neuroradiologists (κ = 0.374). The high Jaccard scores among humans (0.644 for clinicians) indicate a high overlap with ground truth decisions across the full range of treatment options.

### LLM Diagnostic Accuracy evaluation

Accuracy and Jaccard index values for MT and IVT decisions are summarized in Table 2. Among LLMs, DeepSeek R1 demonstrated the best MT accuracy at 0.87 (95% CI, 0.80– 0.94) and the highest Jaccard index (0.512), followed by OpenAI o3 mini (MT accuracy 0.80; Jaccard 0.473). Gemini 2.0 and LLaMA 3.3 showed lower IVT accuracy (0.65 and 0.63, respectively) and comparatively lower Jaccard scores (0.347 and 0.285). Across both human and LLM groups, MT accuracies were generally higher than IVT accuracies, reflecting the increased complexity and variability of IVT decision-making. Figure 1 displays the accuracy of LLMs and clinicians.

**Table 2.** Accuracies for MT, IVT and Jaccard Index.

| Model | Accuracy for MT (95% CI) | Accuracy for IVT (95% CI) | Jaccard Index |
| --- | --- | --- | --- |
| Neurologists | 0.81 (0.71 - 0.88) | 0.80 (0.70 - 0.87) | 0.619 |
| Neuroradiologists | 0.84 (0.74 - 0.90) | 0.76 (0.66 - 0.84) | 0.538 |
| Clinicians (all) | 0.84 (0.74 - 0.90) | 0.80 (0.70 - 0.87) | 0.644 |
| DeepSeek R1 | 0.87 (0.80 - 0.94) | 0.78 (0.69 - 0.87) | 0.512 |
| Gemini 2.0 | 0.81 (0.73 - 0.89) | 0.65 (0.55 - 0.75) | 0.347 |
| LLaMA 3.3 | 0.76 (0.67 - 0.86) | 0.63 (0.52 - 0.74) | 0.285 |
| OpenAI o3 mini | 0.80 (0.71 - 0.89) | 0.76 (0.67 - 0.86) | 0.473 |

**Figure 1.**
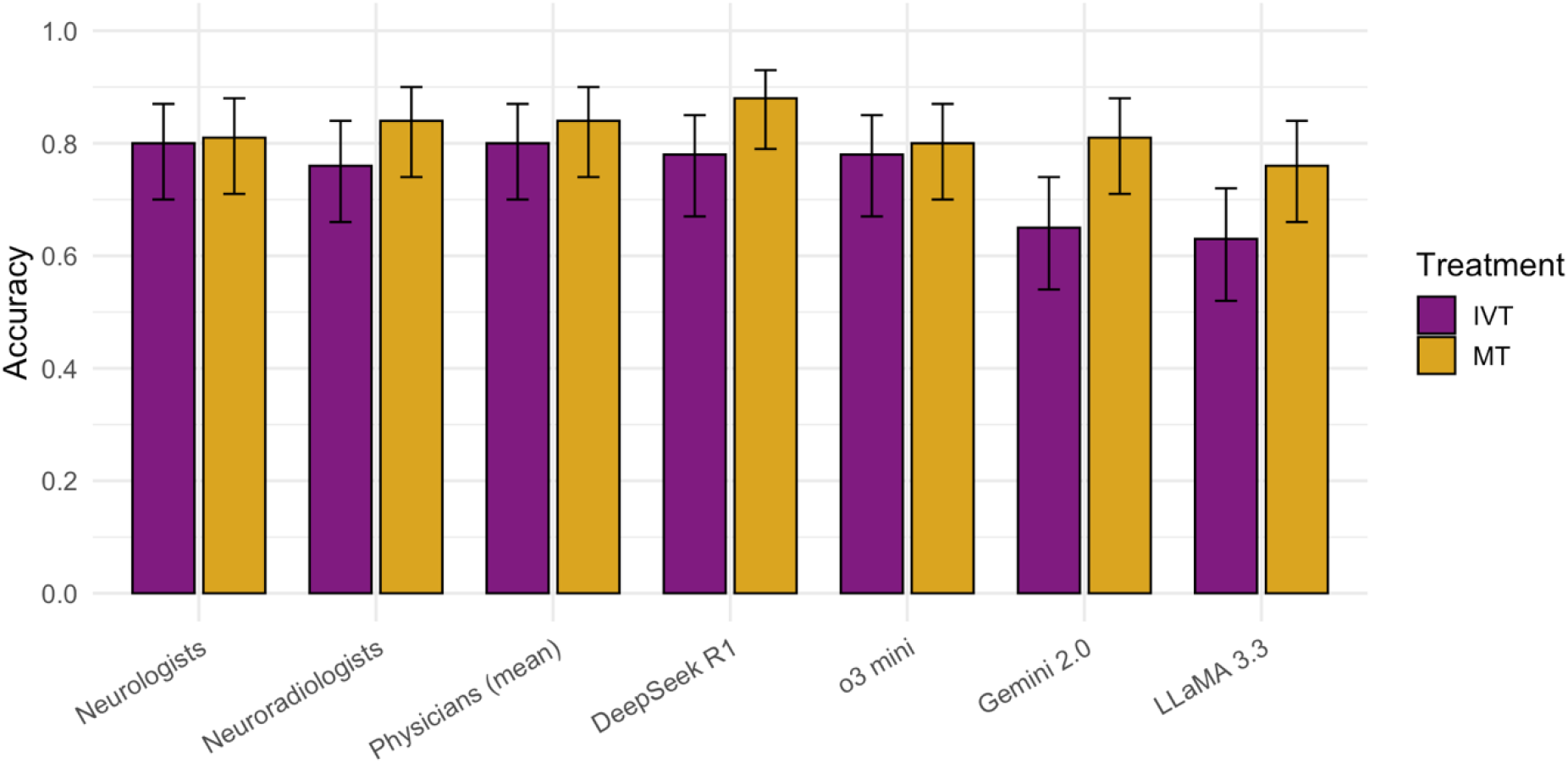
Accuracy of Large Language Models and Stroke Experts in Acute Ischemic Stroke Treatment Decisions. Bar plot showing the accuracy (mean ± 95% confidence interval) of four large language models (DeepSeek R1, OpenAI o3 mini, Gemini 2.0, LLaMA 3.3) and stroke experts (neurologists, neuroradiologists, and pooled physicians) in recommending intravenous thrombolysis (IVT, purple) and mechanical thrombectomy (MT, yellow) for 80 acute ischemic stroke cases.

### LLM Reasoning Evaluation

Error composition analysis revealed distinct patterns across large language models (LLMs) and treatment decision categories (Figure 2). In the *Mechanical Thrombectomy* (MT) group, most cases were free from reasoning errors, ranging from 79% for LLaMA 3.3 to 89% for DeepSeek R1. The most frequent error category in MT was *Discrepant Ground Truth*, accounting for 8–15% of cases depending on the model, followed by *Borderline Case Misclassification* (1–5%). *Guideline non-adherence* was rare (<5%), and no omissions of critical information were observed in any MT decision. Detailed error rates for MT across categories and LLMs are presented in Table 3.

**Table 3.** Errors in Mechanical Thrombectomy group (n=80)

| **LLM** | **Discrepant Ground Truth** | **Guideline non-adherence** | **Omission of Critical Information** | **Borderline Case Misclassification** |
| --- | --- | --- | --- | --- |
| OpenAI o3 mini | 12 (15%) | 1 (1%) | 0 (0%) | 2 (3%) |
| Gemini 2.0 | 10 (13%) | 2 (3%) | 0 (0%) | 1 (1%) |
| LLaMA 3.3 | 9 (11%) | 4 (5%) | 0 (0%) | 4 (5%) |
| DeepSeek R1 | 6 (8%) | 0 (0%) | 0 (0%) | 2 (3%) |

**Figure 2.**
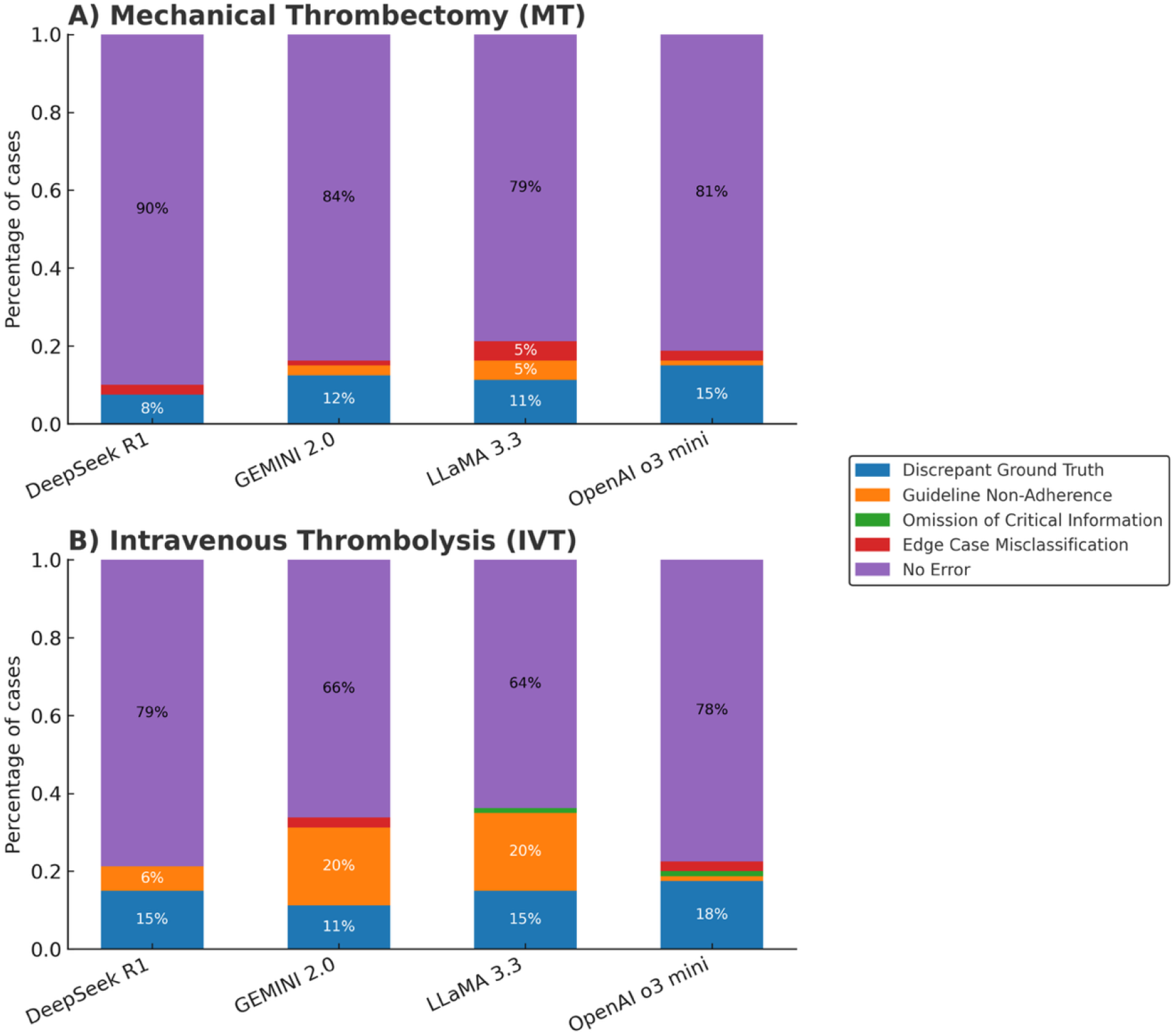
Error composition of LLMs in treatment decision-making for acute ischemic stroke. Stacked bar plots showing the proportion of cases with different reasoning error types across four large language models for (A) Mechanical Thrombectomy (MT) and (B) Intravenous Thrombolysis (IVT).

In contrast, the *Intravenous Thrombolysis* (IVT) group demonstrated slightly higher proportions of reasoning errors, with “error-free” rates between 64% (LLaMA 3.3) and 79% (DeepSeek R1). *Discrepant Ground Truth* errors occurred in 11–18% of IVT cases, while *Guideline non-adherence* was notably more frequent, reaching 20% for GEMINI 2.0 and LLaMA 3.3. *Omission of Critical Information* was rare (<2%), and *Edge Case Misclassification* remained ≤3%. Detailed error rates for IVT across categories and LLMs are presented in Table 4.

**Table 4.**
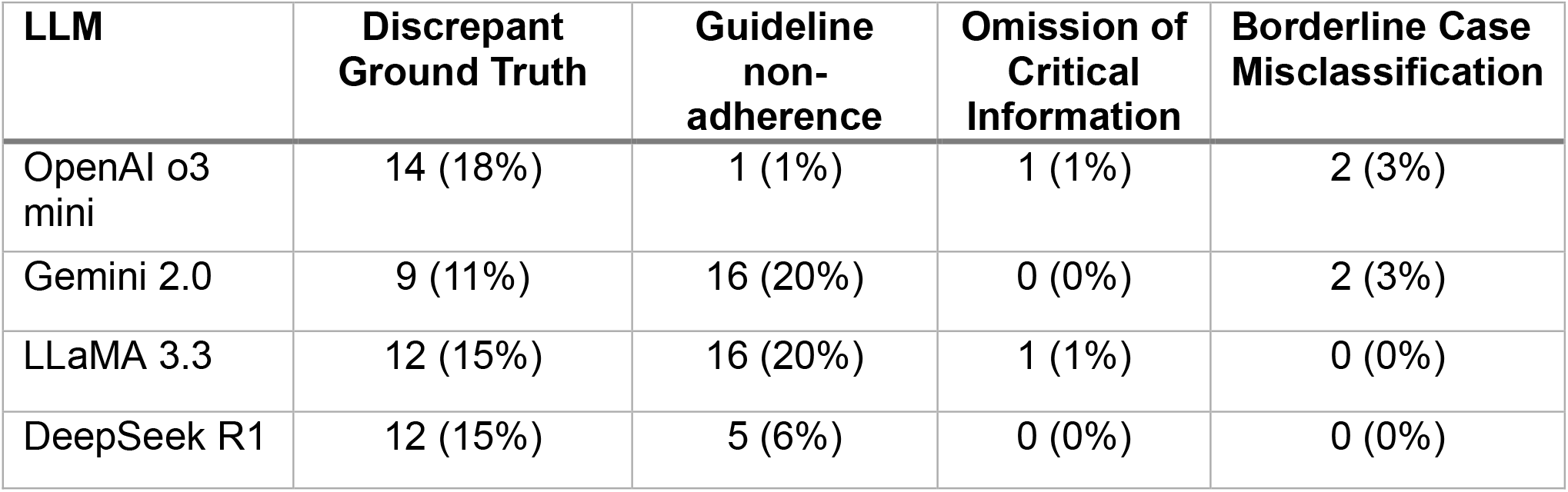
Errors in Intravenous Thrombolysis group (n=80)

| **LLM** | **Discrepant Ground Truth** | **Guideline non-adherence** | **Omission of Critical Information** | **Borderline Case Misclassification** |
| --- | --- | --- | --- | --- |
| OpenAI o3 mini | 14 (18%) | 1 (1%) | 1 (1%) | 2 (3%) |
| Gemini 2.0 | 9 (11%) | 16 (20%) | 0 (0%) | 2 (3%) |
| LLaMA 3.3 | 12 (15%) | 16 (20%) | 1 (1%) | 0 (0%) |
| DeepSeek R1 | 12 (15%) | 5 (6%) | 0 (0%) | 0 (0%) |

Notably, two patients were excluded from the error analysis: one declined treatment, and the other was enrolled in a clinical study. The supplementary file provides illustrative examples of LLM-generated reasoning across different error categories.

## Discussion

Acute ischemic stroke (AIS) management is a complex, multidisciplinary process requiring coordinated input from neurologists, neuroradiologists, anesthesiologists, and well-equipped stroke units [6]. Despite advances in regional stroke networks, approximately one third of stroke patients initially presents to community hospitals and must be transferred to comprehensive stroke centers for definitive treatment [7]. In such settings, the lack of immediate specialist expertise may delay critical decisions, potentially jeopardizing patient outcomes [20].

Our findings suggest that large language models (LLMs) with advanced reasoning capabilities could serve as effective decision support tools in this context. Their performance in determining eligibility for mechanical thrombectomy (MT) and intravenous thrombolysis (IVT) matched or even exceeded that of experienced physicians. Notably, the top-performing model, DeepSeek-R1, achieved an MT decision accuracy of 87%, outperforming neurologists (81%) and neuroradiologists (84%). These results highlight the potential of generative AI to support high-stakes, time-critical decisions in stroke care, particularly in scenarios where immediate specialist consultation is not available.

In our retrospective, multi-center study, 80 real-world cases from two large stroke centers in different European countries were evaluated. Fourteen clinical variables were provided to both human raters and LLMs, including demographic, clinical, and imaging data. Neurologists (n=2) achieved accuracies of 0.81 for MT and 0.80 for IVT, while neuroradiologists (n=4) achieved 0.84 for MT and 0.76 for IVT. For MT decisions, interrater agreement was moderate among neurologists (κ = 0.618) and neuroradiologists (κ = 0.595), indicating reasonable consistency. IVT decisions showed substantial agreement among neurologists (κ = 0.725) but only fair agreement among neuroradiologists (κ = 0.374), reflecting differences in clinical roles; neurologists typically lead IVT decision-making, whereas neuroradiologists focus primarily on MT eligibility.

The LLMs demonstrated robust performance in integrating heterogeneous clinical inputs and applying relevant treatment guidelines. DeepSeek-R1, an open-source model, achieved 87% accuracy for MT and 78% for IVT. By contrast, LLaMA 3.3 ranked lowest, with 76% for MT and 63% for IVT. These results indicate that both commercial and open-source LLMs can synthesize demographic, clinical, and imaging information to generate patient-specific treatment recommendations, with some open-source models performing comparably to proprietary systems, an important consideration for privacy-preserving clinical deployment.

The consistently higher accuracies observed for MT compared to IVT across both human raters and LLMs likely reflect differences in decision complexity. MT eligibility is primarily determined by well-defined imaging and clinical criteria, such as large vessel occlusion and time from symptom onset, which may be more straightforward to assess. In contrast, IVT decisions require consideration of a broader range of clinical factors, including contraindications (e.g., recent surgery, anticoagulation status, bleeding risk) and time-sensitive nuances, which introduce variability and uncertainty. This complexity may explain why IVT accuracy was more variable and generally lower, particularly among LLMs, and why guideline non-adherence errors were more prevalent in the IVT reasoning evaluation.

Our results are in line with Kottlors et al., who investigated GPT-3 for thrombectomy triage using radiology reports and limited clinical data in a single-center German cohort [21]. In their study, the LLMs achieved an overall accuracy of 88%, with a specificity of 96% and a sensitivity of 80%. The present study extends these findings by incorporating a more heterogeneous, multi-center dataset and benchmarking performance against both expert consensus and real-world treatment decisions. Moreover, in addition to the binary “yes/no” outputs in Kottlors et al., our LLMs were required to provide explanatory rationales for their recommendations. This feature not only improves transparency and clinician trust but also offers educational value by making the decision-making process explicit. Additionally, we assessed both commercial and open-source models, demonstrating that open-source systems can achieve non-inferior performance without the data privacy concerns associated with proprietary cloud-based tools.

The ability of LLMs to produce reasoning is particularly important in clinical AI applications. Even accurate recommendations may be dismissed by clinicians if not accompanied by an explanation. Our error composition analysis revealed clear differences between treatment decision types. For MT, most LLM outputs were free from reasoning errors (79–89%), with Discrepant Ground Truth errors being the most common (8–15%), followed by infrequent Borderline Case Misclassifications (1–5%) and rare guideline deviations (<5%). No omissions of critical information occurred.

In contrast, IVT decisions showed lower rates of error-free reasoning (64–79%) and a higher incidence of Guideline non-adherence, particularly in Gemini 2.0 and LLaMA 3.3 (20%). Discrepant Ground Truth Errors were observed in 11–18% of cases, while omissions of critical information (<2%) and borderline case misclassifications (≤3%) remained uncommon. These findings indicate that, while all models maintained a high proportion of error-free reasoning in MT decisions, IVT recommendations were more susceptible to errors, reflecting the greater complexity of IVT decision-making, which involves multiple eligibility criteria, time-dependent factors, and considering numerous contraindications.

This study has several limitations. Although based on real-world cases from two centers, the ground truth remains subject to retrospective bias and variability in interdisciplinary decision-making. Some influencing factors, such as family discussions or pre-defined treatment limitations, were not documented and therefore not available to neither LLMs nor human raters. Additionally, while we evaluated several state-of-the-art LLMs, performance was not uniform, and generalizability to other populations and health systems remains uncertain. Finally, while we assessed reasoning quality qualitatively, more standardized and quantitative metrics for evaluating factual accuracy, logical coherence, and guideline adherence in AI-generated rationales are needed.

In conclusion, our study demonstrates that LLMs can match or even surpass expert clinician performance in AIS treatment eligibility decisions, while also providing explainable rationales that may enhance clinician trust and facilitate learning. Compared with prior work, our approach leverages a more complex, real-world dataset, incorporates reasoning-based evaluation, and highlights the viability of open-source models for privacy-preserving deployment. Future research should focus on prospective, real-time validation in clinical workflows, multilingual adaptability, and integration with stroke imaging and workflow data to create robust, trustworthy decision support systems for acute stroke care.

## Data Availability

The datasets analyzed during the current study are not publicly available due to patient privacy and institutional data protection regulations. Upon acceptance of this manuscript in a peer-reviewed journal, the code used for data analysis and model evaluation will be made available in a publicly accessible GitHub repository.

